# Divergences on expected pneumonia cases during the COVID-19 epidemic in Catalonia: A time-series analysis of primary care electronic health records covering about 6 million people

**DOI:** 10.1101/2020.12.31.20249076

**Authors:** Ermengol Coma, Leonardo Méndez-Boo, Núria Mora, Carolina Guiriguet, Mència Benítez, Francesc Fina, Mireia Fàbregas, Elisabet Balló, Francisa Ramos, Manuel Medina, Josep M. Argimon

## Abstract

**Background:** Pneumonia is one of the complications of COVID-19. Primary care electronic health records (EHR) have shown the utility as a surveillance system.

**Aim:** To analyze the trends of pneumonia during two waves of COVID-19 pandemic in order to use it as a clinical surveillance system and an early indicator of severity.

**Methods:** Time series analysis of pneumonia cases, January 2014-December 2020. We collected pneumonia diagnoses from primary care EHR, covering >6 million people in Catalonia (Spain). We compared the trend of pneumonia in the season 2019-2020 with that in the previous years. We estimated the expected pneumonia cases with data from 2014 to 2018 using a time series regression adjusted by seasonality and influenza epidemics.

**Results:** Between 4 March and 5 May 2020, 11,704 excess pneumonia cases (95% CI: 9,909 to 13,498) were identified. We observed a second excess pneumonia period from 22 october to 15 november of 1,377 excess cases (95% CI: 665 to 2,089). In contrast, we observed two great periods with reductions of pneumonia cases in children, accounting for 131 days and 3,534 less pneumonia cases (95% CI: 1,005 to 6,064) from March to July; and 54 days and 1,960 less pneumonia cases (95% CI 917 to 3,002) from October to December.

**Conclusions:** Diagnoses of pneumonia from the EHR could be used as an early and low cost surveillance system to monitor the spread of COVID-19.

## Introduction

The coronavirus disease 2019 (COVID-19) caused by the severe acute respiratory syndrome coronavirus 2 (SARS-CoV-2) started as an outbreak in Wuhan (China) and rapidly evolved into a worldwide pandemic and a public health emergency of international concern [1]. As of 11 december 2020, more than 69 million people have contracted the virus worldwide, with more than 1.5 million confirmed deaths [2]. Asymptomatics seem to account for approximately 20% of infected [3]. Most cases present mild influenza-like illness symptoms including fever, cough, sore throat, fatigue and myalgia [4]. Around 10-20% of symptomatic patients present severe forms of disease that require hospital admission, mainly due to pneumonia with severe inflammation [4 - 6]. Lung damage can progress rapidly and an early detection is essential for better management and also for surveillance of medium and long-term sequelae [7].

The first case in Europe was reported on 20 January 2020 in France and the first case in Catalonia (Spain) was on 25 February 2020. However, some studies have suggested a previous circulation of the virus [8, 9]. Due to the rapid spread of the disease, on March 14 Spain established a national lockdown, as many other countries [10]. During the first surge, as Spain lacked the capacity to test all cases, reverse transcriptase–polymerase chain reaction (RT-PCR) test confirmation was only required when patients were admitted to hospital or were healthcare staff. In a previous study, only 38.5% of clinical COVID-19 cases diagnosed in primary care between March 1 and April 24 2020 received a RT-PCR test [5]. On May 11, primary care acquired the capacity to perform RT-PCR test and the number of tests increased as stated in the official figures of the Catalan Health Department website (https://dadescovid.cat/).

Knowledge of the infection status and continuous measurement of the transmission is one of the five necessary components described for containing COVID-19 [11].This measurement should also include surveillance based on clinical diagnoses. Primary care clinical diagnoses have been used as a surveillance system for influenza epidemics during many years [12]. In February and March 2020, we found an excess of influenza diagnoses using this clinical surveillance system that suggested an early spread of COVID-19 in Catalonia prior to the first reported case [9]. This finding showed the utility of flu clinical diagnoses as a potential low-cost surveillance system capable of early monitoring the spread of the epidemic. However, as flu cases are limited to winter, we need comprehensive and complementary information to understand the actual trend of the COVID-19 epidemic, including severity.

In addition to the current system, the aim of our study is to analyze pneumonia diagnoses and compare the trend in the 2019-2020 season with those of the previous years in order to add another surveillance measurement of the evolution of the epidemic that could add a severity component.

## Methods

We performed a time-series study of clinical diagnoses of pneumonia. The study period included 6 years from 1 September 2014 to 4 december 2020 considered as seasons from autumn to summer as routinely done for influenza epidemics.

Our main outcome was clinical diagnoses of pneumonia. Daily counts of diagnoses of pneumonia were retrospectively extracted from the primary care electronic health records (EHR) of the Catalan Institute of Health (ICS for its Catalan initials). ICS is the main health provider in Catalonia. It manages about 75% of all primary care practices in the Catalan public health system and covers about 6 million people [13]. All general practices use the same EHR known as ECAP. The ECAP is a software system that serves as a repository for structured data on diagnoses (coded according to the International Classification of Diseases 10th revision ICD-10), clinical variables, prescription data, laboratory test results, visits and diagnostic requests.

We defined pneumonia diagnoses according to the ICD-10 classification and included all codes presented in supplementary table 1. COVID-19 confirmed cases were obtained from the official website of the Catalan Health department Dadescovid.cat (https://dadescovid.cat/).

**Table 1.** Periods and number of excess pneumonia cases in Catalonia, percentage of excess pneumonia compared to the expected and the number of days with excess within the period from 1 September 2019 to 4 December 2020.

| Age group* | Period** of excess pneumonia cases | Estimated number of excess pneumonia cases (95% CI) | Percentage compared to expected cases (95% CI) | Number of days with excess cases within the period |
| --- | --- | --- | --- | --- |
| Between 15 and 64 | 16/01/2020 - 19/01/2020 | 53<br>(7 - 100) | 22.69<br>(2.41 - 52.97) | 4 |
|  | 27/01/2020 - 25/02/2020 | 587<br>(238 - 935) | 34.29<br>(11.56 - 68.65) | 30 |
|  | 02/03/2020 - 12/05/2020 | 8,274<br>(7,4371 - 9,110) | 279.9<br>(196.1 - 430) | 72 |
|  | 13/08/2020 - 16/08/2020 | 54<br>(8 - 101) | 77.23<br>(6.83 - 419.7) | 4 |
|  | 19/10/2020 - 24/11/2020 | 1562<br>(1,132 - 1,991) | 130<br>(69.36 - 258.1) | 37 |
| Older than 64 | 05/12/2019 - 16/12/2019 | 165<br>(61 - 269) | 38.61<br>(11.53 - 83.05) | 12 |
|  | 29/12/2019 - 11/01/2020 | 195<br>(74 - 317) | 29.34<br>(9.41 - 58.16) | 14 |
|  | 28/01/2020 - 28/02/2020 | 425<br>(149 - 702) | 27.21<br>(8.08 - 54.57) | 32 |
|  | 02/03/2020 - 13/05/2020 | 5,010<br>(4,378 - 5,641) | 169.2<br>(121.9 - 242.1) | 73 |
|  | 17/08/2020 - 26/08/2020 | 115<br>(29 - 202) | 61.62<br>(10.59 - 200.1) | 10 |
|  | 18/09/2020 - 23/09/2020 | 63<br>(11 - 115) | 45.13<br>(5.64 - 131.8) | 6 |
|  | 02/10/2020 - 08/10/2020 | 74<br>(13 - 134) | 38.67<br>(5.19 - 103.4) | 7 |
|  | 19/10/2020 -<br>29/11/2020 | 1,147<br>(783 - 1,510) | 87.45<br>(46.79 - 159.3) | 42 |
| Total | 08/12/2019 -<br>14/12/2019 | 273<br>(74 - 473) | 30.18<br>(6.68 - 66.97) | 7 |
|  | 28/01/2020 -<br>02/02/2020 | 198<br>(28 - 369) | 19.94<br>(2.36 - 44.81) | 6 |
|  | 04/02/2020 -<br>19/02/2020 | 555<br>(99 - 1,011) | 20.9<br>(3.18 - 45.97) | 16 |
|  | 04/03/2020 -<br>05/05/2020 | 11,704<br>(9,909 - 13,498) | 150.2<br>(103.4 - 225.1) | 63 |
|  | 22/10/2020 -<br>15/11/2020 | 1,377<br>(665 - 2,089) | 57.01<br>(21.27 - 122.6) | 25 |

\*Population younger than 15 years didn't show any excess pneumonia cases during the study period.
\*\*Periods were only considered if excess cases extends more than 3 days

### Statistical analysis

Daily counts of pneumonia cases were computed based on the frequency of cases recorded in the previous 7-day period to avoid weekly effects on recording practice.

We obtained the expected cases for the study period using a time series regression adjusted by seasonality and influenza epidemics. Flu cases were also computed as daily counts in a 7-day period as done for pneumonia and COVID-19 cases and were extracted from the same EHR.

Dataset was divided into three sets: training set (from September 2014 to August 2018), validation set (from September 2018 to August 2019) and analysis set (from September 2019 to December 2020). We used the training set to adjust the model and validation set to test our method as a sensitivity analysis. We checked whether our method identified any excess or lack of pneumonia cases in a regular year not affected by the COVID-19 pandemic. Finally, we projected the estimated time series to our analysis set.

The expected cases were estimated using data from the training seasons adjusted for the influenza epidemics. Excess pneumonia cases were defined as the number of observed minus the expected cases in all periods where observed cases were greater than the upper 95% confidence interval (95% CI). Similarly, the lack or reduction of pneumonia cases was defined as the difference between the expected minus the observed for all periods in which the observed number was below the lower 95% CI. Excess and lack of pneumonia were only calculated for the analysis set (2019-2020 season).

Time-series analysis was performed globally and for age groups (< 15 years old, 15-64 years old, >64 years old). We calculated 95% CIs for each estimate.

All analyses were performed in R v.3.5.1 [14]. All data and analytical code are provided at https://github.com/ErmengolComa/pneumonia.git

### Ethical statement

This study was done in accordance with existing statutory and ethical approvals from the Clinical Research Ethics Committee of the IDIAPJGol (project code: 20/172-PCV).

## Results

Between 1 September 2014 and 4 December 2020 we observed 260,910 pneumonia cases of whom 28.7% were diagnosed in the population younger than 15 years, 37.1% in the population between 15 and 64 years and 34.2% in the population older than 64 years. The mean number of pneumonia diagnoses was 40,224 for seasons between 2014-2015 and 2018-2019. The 2019-2020 season included 50,039 cases, an increase of 24.4%.

In five out of six seasons included in our study, the peak of pneumonia cases coincided with the peak of the influenza epidemics, except for the season 2019 - 2020 (figure 1).

**Figure 1.**
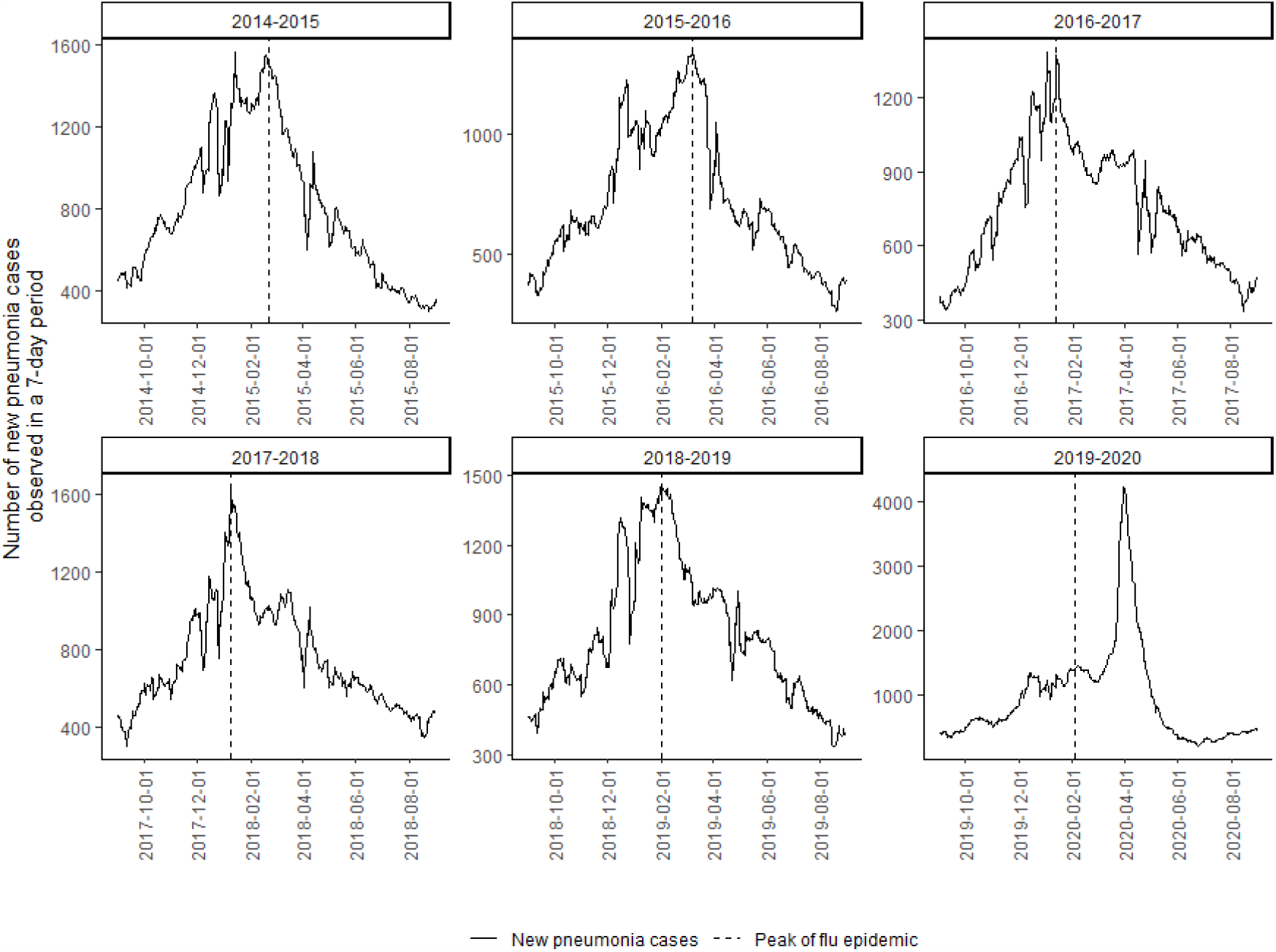
Number of new pneumonia cases observed in a 7-day period by season.

Figure 2 shows the observed and estimated number of 7-day new pneumonia cases (with 95% CI) by age groups, for the validation and the analysis sets. In the whole population, we didn’t observe large periods of excess or lack of pneumonia during the validation set (2018-2019). Nonetheless, between 4 March and 5 May 2020, we observed a great period of excess pneumonia cases, accounting for 63 days and 11,704 excess cases (95% CI: 9,909 to 13,498). There were other periods of excess pneumonia before, during December 2019, late January 2020 and early February 2020, although they account for less days and less cases. In addition, 25 days of excess were observed from 22 October 2020 to 15 November 2020, accounting for 1,377 excess cases (95%CI 665 - 2,089) (Table 1).

**Figure 2.**
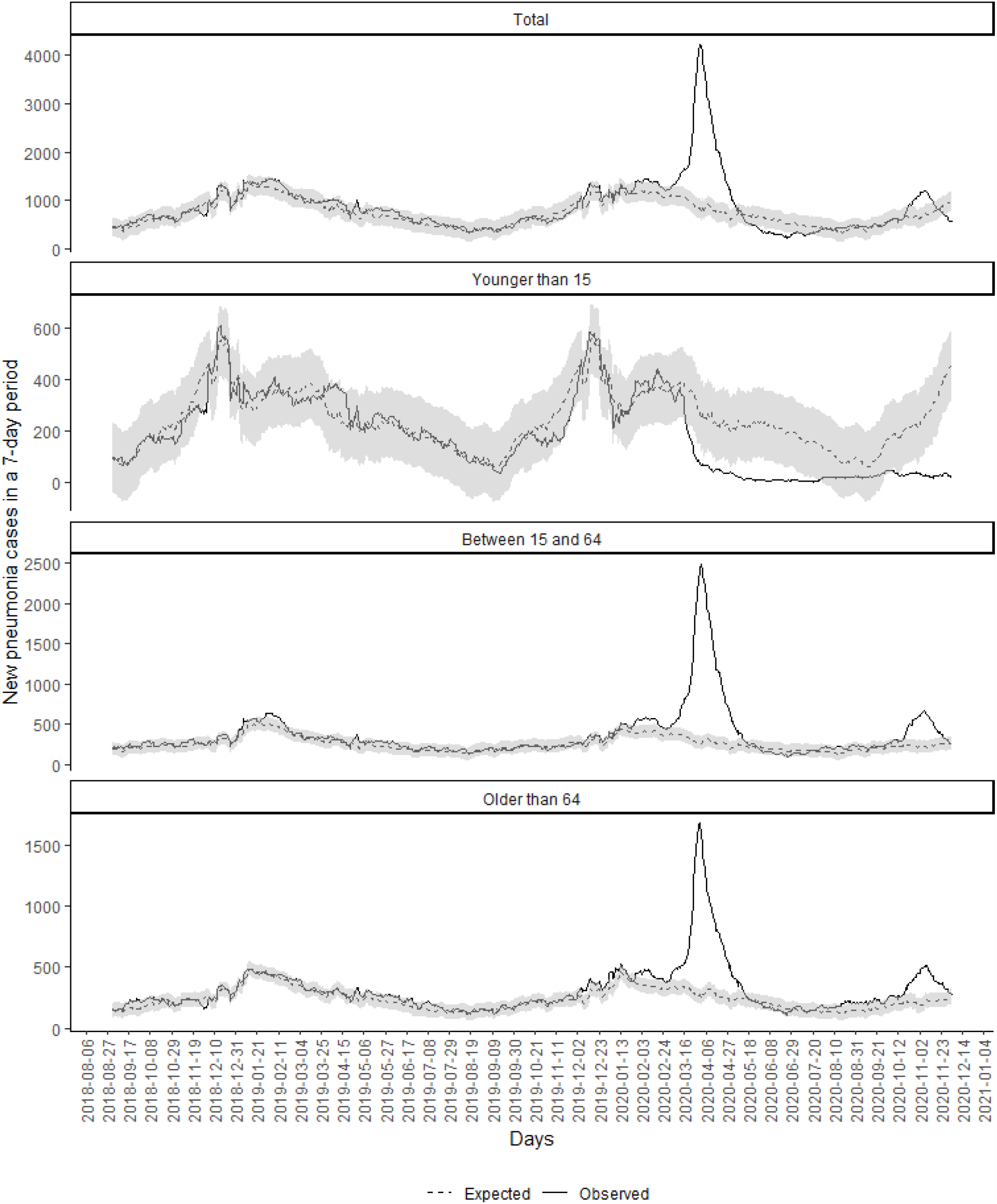
Observed and expected (with 95% CI) new pneumonia cases in a 7-day period from September 2018 to December 2020 in Catalonia.

Table 1 also presents the different periods with excess pneumonia cases stratified by age group. Population between 15 and 64 years and the population older than 64 had similar periods of excess cases, accounting for 72 and 73 days and 8,274 and 5,010 cases during the March-May excess, respectively. And accounting for 37 days and 1,562 cases and 42 days and 1,147 cases respectively during the October-November excess period. Conversely in children we didn’t observe any excess during the whole period of the study (Figure 2 and Table 1).

In contrast, regarding the lack of pneumonia, we observed a reduction of pneumonia cases among people younger than 15 years between 18 March and 26 July, accounting for 131 consecutive days with less observed pneumonia than expected, 3,534 less observed cases during that period (95% CI: 1,005 to 6,064) and a reduction of 84.43% (95% CI 60.67% to 90.3%) compared with the expected. In addition, another period of lack of pneumonia was observed in children from 12 October to 4 December, with 54 days of reductions and a 1,960 less cases (95% CI 917 - 3002).We didn’t observe any period of reduction of pneumonia for other age groups in the analysis set (Table 2).

**Table 2.** Periods and number of reductions of pneumonia cases in Catalonia, the percentage of reductions compared to the expected and the number of days with reduction within the period from 1 September 2018 to 4 December 2020.

| Age group* | Period** of<br>reduction of<br>pneumonia<br>cases | Estimated number<br>of reduction of<br>pneumonia cases<br>(95% CI) | Percentage<br>compared to<br>expected cases<br>(95% CI) | Number of days<br>with reduction of<br>pneumonia cases<br>within the period |
| --- | --- | --- | --- | --- |
| Younger than 15 | 18/03/2020 -<br>26/07/2020 | 3,534<br>(1,005 - 6,064) | 84.43<br>(60.67 - 90.30) | 131 |
|  | 06/10/2020 -<br>10/10/2020 | 104<br>(7 - 200) | 77.04<br>(18.45 - 86.64) | 5 |
|  | 12/10/2020 -<br>4/12/2020 | 1,960<br>(917 - 3,002) | 89.15<br>(79.37 - 92.64) | 54 |

\*We presented only data from the population younger than 15 years because we didn't find any reductions in other age groups.
\*\*Periods were only considered if reduction of cases extends more than 3 days

Finally, Figure 3 shows the trend of pneumonia and COVID-19 cases from March 2020. We observed two waves of COVID-19 cases coinciding with two pneumonia peaks in population older than 15 years.

**Figure 3.**
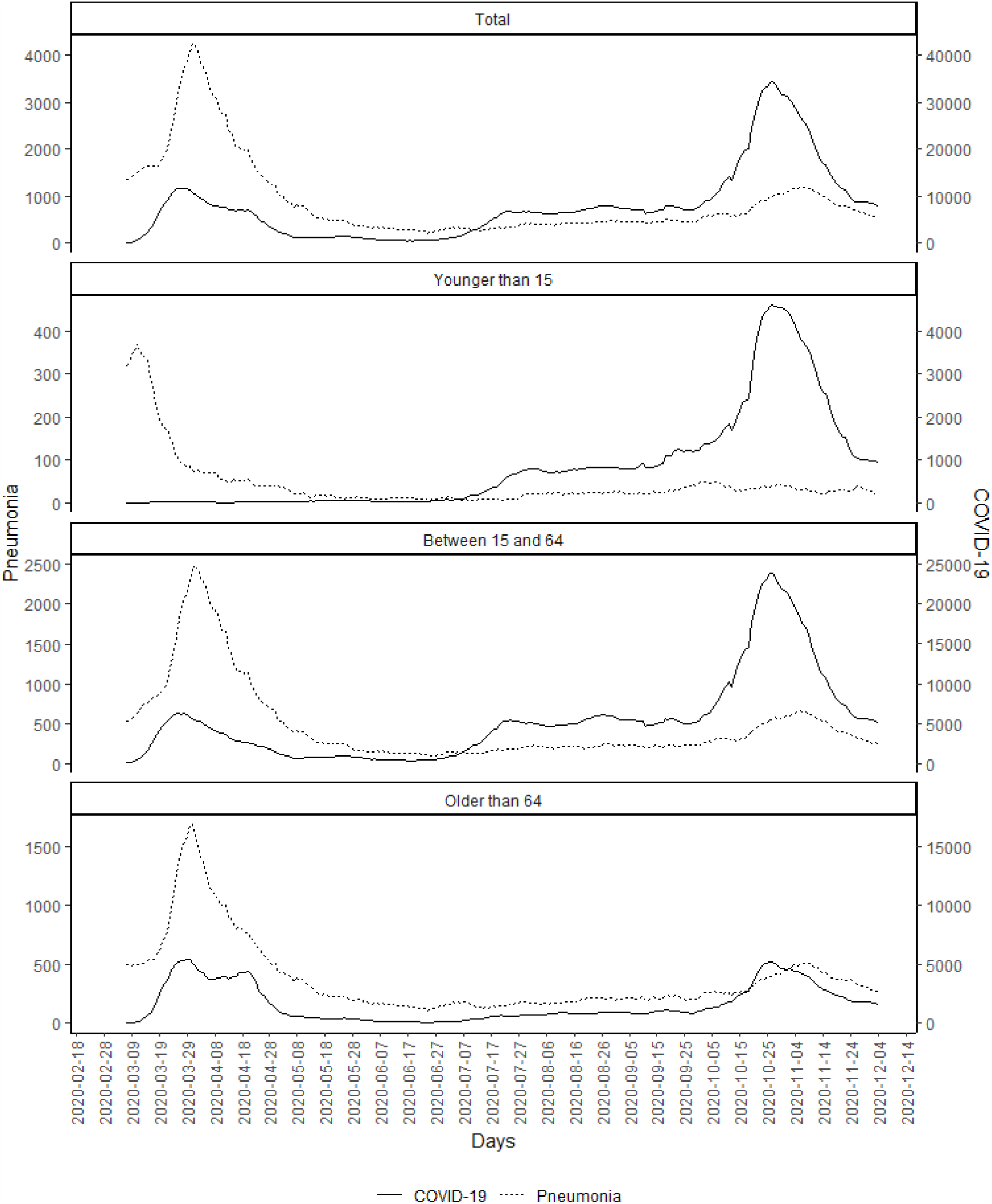
Number of new pneumonia and COVID-19 cases in the previous 7-day, from 1st March 2020.

## Discussion

In our work, we found an excess of 11,704 pneumonia diagnoses between 4 March 2020 and 5 May 2020. This excess seems to be related to the COVID-19 pandemic, as temporally overlapped with the first wave in Spain. The excess found only a few days after the first official case of COVID-19 in Catalonia also suggests that SARS-CoV-2 could have been circulating in the Catalan population when the first imported case was reported on 25 february 2020 [9]. Prior to this large excess, we found a small excess period in mid-December that doesn’t seem related to COVID-19 as it only lasted for 7 days and then observed cases went down to the expected for at least one month. But after that, it is remarkable that between 28 January and 19 February 2020 (one month before the first confirmed COVID-19 case was reported in Catalonia), we found 22 out of 23 days of excess of pneumonia separated into two periods that should be accounted as the same. Although this period matches with the peak of influenza epidemic, it could also be related to SARS-CoV-2 transmission as it occurred only 15 days before the huge excess described above. This hypothesis was launched in a previous study where we found an excess of influenza cases from the beginning of February in Catalonia [9] and was also suggested in other studies in different countries [8]. In addition, a CDC report estimated that the start of SARS-CoV-2 communitary transmission was around mid January to February [15]. Finally, scientists found virus traces in wastewater collected in January in Barcelona [16] and in December in Italy [17]. Our results, then, are in line with those published until now and strengthen this hypothesis.

After May, we didn’t find any other excess until mid October, coinciding with the second wave of the COVID-19 pandemic in Catalonia. However, this excess was lesser than the one found in the March-May period, suggesting a lower severity of the situation. Official data from Spanish government about excess mortality shows an excess of 120% in Catalonia from 13th March to 8th May and an excess of 25.8% from 6th October to 28th November. So, similar to our findings in pneumonia, severity of the second wave was lesser than the first one and pneumonia advanced some days as an alert of increased severity, without delays as occurred in mortality notifications. Nonetheless, the number of COVID-19 cases was greater because testing increased since May in Catalonia. Using pneumonia as a surveillance indicator could help to understand the severity of the wave regardless of the capacity to test.

Trends of pneumonia in children followed a different pattern. We found a greater lack of pneumonia cases during the lockdown in patients younger than 15 years old. This is an expected finding, as schools closed on March 13 and didn’t reopen until September 14. In addition, mobility measures during the lockdown were stricter in children, while adults were able to move for essential work or to shop. This reduction was also found in other infectious diseases. For example, in Diagnosticat (https://www.ics.gencat.cat/sisap/diagnosticat/principal), an official website of the Catalan Health Department that publishes weekly clinical diagnoses of 7 notifiable infectious diseases since 2010 [12], we saw a decrease in the weekly rates of chickenpox from lockdown onset as it happens with pneumonia. But most importantly, we didn’t find any excess pneumonia in patients younger than 15 years during February and early March, before the lockdown. This seems to suggest that COVID-19 is less severe in children, as other studies have pointed out [19]. More surprising are the results found in October-December. Despite schools reopened in September in Catalonia (with measures), diagnoses of pneumonia in children continued below the expected. Several studies have observed low transmission and severity of COVID-19 in educational settings, which could explain our findings [19]. In addition, this reduction of pneumonia in children also suggests that other common pathogens that cause pneumonia in children could have low transmission possibly due to anti-covid measures [20]. This is consistent with the data of the southern hemisphere. In Australia, researchers observed reductions of 99% of respiratory syncytial virus (RSV) infections during its winter that they considered related to COVID-19 control measures [21]. In Catalonia, measures in schools were the use of masks for children older than 6 years, bubble groups, not mixing children from different groups and daily symptoms screening. More studies are needed to confirm if some of these measures affected the transmission of other pathogens or there are other factors that caused a great reduction of all types of pneumonia even with schools open, such as a viral competition between SARS-CoV-2 and other viruses [22]. This could be of interest for the following seasons.

Our research has several limitations. Firstly, the design of our study does not allow us to ensure a causal link between COVID-19 epidemic and the excess of pneumonia, but only a temporal coincidence. Moreover, as we lacked tests we were not able to differentiate the etiology of each pneumonia. However, our method offers a low-cost surveillance system that could help to detect unusual trends, supporting public health responses. Secondly, as our study uses data from several years, changes in population structure could limit the use of this method. Nonetheless, population structure in terms of age and gender has remained stable in the study period [23]. Thirdly, using data from the primary care EHR could introduce some bias as we lacked information about emergency departments or hospital admission.

The strengths of this study include population-based data automatically obtained from primary care EHR. Several studies have used the Catalan EHR to do useful research in real-world conditions and for the surveillance of different diseases [9, 24]. Our database covers over 75% of the population of Catalonia, allowing us to detect general and local excesses and lack of pneumonia. It’s also a quick and low cost method to integrate in the current information systems of any region using EHR. In addition, we have tested our method in a non-COVID-19 affected season (2018-2019) and we didn’t find any unusual pattern, strengthening our subsequent findings. Finally, this study presents for the first time an analysis of trends of expected pneumonia during two waves of COVID-19 epidemic. The consistency of our results is reaffirmed by the reproducibility during the second wave and the similarity with excess mortality figures [18], suggesting that an increase of pneumonia could be an alert of COVID-19 outbreak and future mortality.

In conclusion, monitoring clinical diagnoses of pneumonia and its comparison with what is expected could help interpreting the epidemiological situation. This surveillance system based on routinely collected data from the EHR could be used as an early and a low cost warning system, allowing to advance public health response independently of the capacity to test. In addition, more studies are needed to understand the causes of the reduction of pneumonias in children during COVID-19 epidemic.

## Data Availability

All data and analytical code are provided at https://github.com/ErmengolComa/pneumonia.git

https://github.com/ErmengolComa/pneumonia.git

## Acknowledgments

The authors of this paper would like to thank all primary care professionals in Catalonia for their work and resilience providing care to the catalan population during these difficult times. We also would like to acknowledge the efforts of all members of the SISAP team during the last months.

## Authors’ contributions

All authors contributed to the design of the study, the interpretation of the results, and reviewed the manuscript. EC, FF and NM had access to the data, performed the statistical analysis and acted as guarantors. EC, LM, CG, MB and NM wrote the first draft of the manuscript. All authors critically revised the manuscript. The corresponding author attests that all listed authors meet authorship criteria and that no others meeting the criteria have been omitted.

