## Supplementary Table 1 for "Divergences on expected pneumonia cases during the COVID-19 epidemic in Catalonia: A time-series analysis of primary care electronic health records covering about 6 million people"

**Supplementary Table 1. Pneumonia ICD-10 codes**

| PS_COD | PS_DES |
| --- | --- |
| J11.00 | Influenza due to unidentified influenza virus with unspecified type of pneumonia |
| J12.0 | Adenoviral pneumonia |
| J12.1 | Respiratory syncytial virus pneumonia |
| J12.2 | Parainfluenza virus pneumonia |
| J12.3 | Human metapneumovirus pneumonia |
| J12.81 | Pneumonia due to SARS-associated coronavirus |
| J12.89 | Other viral pneumonia |
| J12.9 | Viral pneumonia, unspecified |
| J13 | Pneumonia due to Streptococcus pneumoniae |
| J14 | Pneumonia due to Hemophilus influenzae |
| J15.1 | Pneumonia due to Pseudomonas |
| J15.211 | Pneumonia due to Methicillin susceptible Staphylococcus aureus |
| J15.29 | Pneumonia due to other staphylococcus |
| J15.4 | Pneumonia due to other streptococci |
| J15.6 | Pneumonia due to other Gram-negative bacteria |
| J15.7 | Pneumonia due to Mycoplasma pneumoniae |
| J15.8 | Pneumonia due to other specified bacteria |
| J15.9 | Unspecified bacterial pneumonia |
| J16.8 | Pneumonia due to other specified infectious organisms |
| J18.0 | Bronchopneumonia, unspecified organism |
| J18.1 | Lobar pneumonia, unspecified organism |
| J18.2 | Hypostatic pneumonia, unspecified organism |
| J18.8 | Other pneumonia, unspecified organism |
| J18.9 | Pneumonia, unspecified organism |
